# Investigating the optimal reactive balance training intensity in people with chronic stroke: study protocol for a randomized control trial

**DOI:** 10.1101/2025.06.26.25330322

**Authors:** Nigel Majoni, Elizabeth L. Inness, David Jagroop, Cynthia J. Danells, Avril Mansfield

**Affiliations:** Toronto Rehabilitation Institute - University Health Network, Toronto, Ontario, Canada; Rehabilitation Sciences Institute, University of Toronto, Toronto, Ontario, Canada; Department of Physical Therapy, University of Toronto, Toronto, Ontario, Canada

## Abstract

Stroke significantly contributes to long-term disability, one of the problems is with impaired balance control, increasing the risk of falls. The risk of falls may be mitigated using reactive balance training (RBT) which has been shown to effectively reduce fall risk by enhancing reactive stepping following repeated balance perturbations. However, the optimal RBT intensity for people with chronic stroke remains unknown. The purpose of this assessor-blinded randomized controlled trial is to investigate the optimal intensity of RBT by comparing high-intensity, moderate-intensity, and walking control groups among 63 individuals with chronic stroke. Participants will undergo four consecutive days of training, with outcomes assessed pre- and post-training and at a one-year follow-up. The primary outcomes are reactive stepping ability, measured using number of steps required to recover balance from a novel perturbation. Secondary outcomes include rates of adverse events, functional balance, falls efficacy, and participation in daily activities. We hypothesize that high-intensity RBT will yield faster adaptations and greater retention compared to moderate-intensity and walking. Determining the optimal RBT intensity could substantially enhance clinical guidelines for stroke rehabilitation, optimize therapy efficiency, and improve patient outcomes by reducing fall risk and improving functional independence. *ClinicalTrials.gov ID NCT06555016*.

## 1. INTRODUCTION

### 1.1 Background and rationale

Stroke is the primary cause of long-term disability.^1,2^ People who have previously experienced a stroke have an increased risk of falling compared to their age-matched counterparts.^3^ Impaired balance control is the main contributor to the increased of falling post-stroke.^4,5^ Balance impairments after stroke negatively affect activities of daily living, increase fear of falling, and decrease quality of life.^6–8^ In individuals who have experienced a stroke, improved balance control predicts return to independent living^9^ and increased quality of life.^10^

Given the importance of balance post-stroke, investigating more effective ways to improve balance among people with stroke warrants further exploration.^10^ Good reactive balance control is essential for preventing falls after a loss of balance.^11^ Reactive balance control can be improved with reactive balance training (RBT). RBT involves participants experiencing repeated balance perturbations to practice and improve the effectiveness of balance reactions.^12^ RBT improves control of reactive stepping in stroke survivors and reduces the rate of falls within various populations, including people with stroke.^13–15^

Despite the benefits of RBT post-stroke, clinicians are uncertain about the optimal training parameters.^16^ The FITT principle is used for prescribing frequency, intensity, type, and time of exercise.^17^ The optimal FITT parameters for RBT in stroke patients remain largely unexplored. Intensity of RBT may be particularly important as intensity and time (duration) of the intervention are often related. That is, high-intensity RBT may improve reactive balance control faster than moderate intensity training, so the same benefits can be achieved in less time.^18^ Among healthy young and older adults, higher perturbation intensities can improve reactive balance control, with improvements of greater step length, and a decrease in the average number of steps required to recover balance.^19,20^ Clinicians have limited time for patient rehabilitation due to competing patient goals and priorities;^16^ therefore, finding training approaches to achieve the same results in less time are crucial. Conversely, high-intensity RBT may increase the risk of adverse events, and people with stroke may have limited tolerance for such intensive interventions. However, no research to date has studied different RBT intensities post-stroke. This research is warranted to clarify the safety, feasibility, and efficacy of high-intensity RBT training in post-stroke.

### 1.2 Objectives and hypotheses

The purpose of this study is to determine the optimal RBT intensity in people with chronic stroke. Our hypotheses are:

1. Adaptation rate will be faster for high-intensity RBT than moderate-intensity RBT;
2. Adaptation rate will be faster for both RBT groups than a walking control group; and
3. Both RBT groups will have better retention of learning than the walking control group; Secondary objectives are to compare the rate of adverse outcomes between groups, and to determine the effect of different intensities of RBT on reactive stepping ability, functional balance, falls efficacy, and participation in daily activities.

### 1.3 Trial design

This an assessor-blinded randomized controlled trial; 63 people with chronic stroke (> 6 months post-stroke) will be randomized into three different intervention groups: 1) high-intensity RBT, 2) moderate-intensity RBT, and 3) a walking control group. We chose a walking control group because we do not expect their performance to improve; therefore, any change reflects the effect of repeated testing only. Training will occur on four consecutive days. At the end of each training session and one week after the last training session, we will assess responses to novel untrained balance perturbations to determine adaptation rate and retention of improvements, respectively. Balance, balance confidence, and quality of life outcomes will be assessed pre-training and one week after the last training session. For 12 months post-training, we will ask participants to track their daily falls, fear of falling, and quality of life (Figure 1). The protocol is reported according to the Standard Protocol Items: Recommendations for Interventional Trials,^21^ Template for Intervention Description and Replication,^22^ and the Consensus on Exercise Reporting Template.^23^

**Figure 1.**
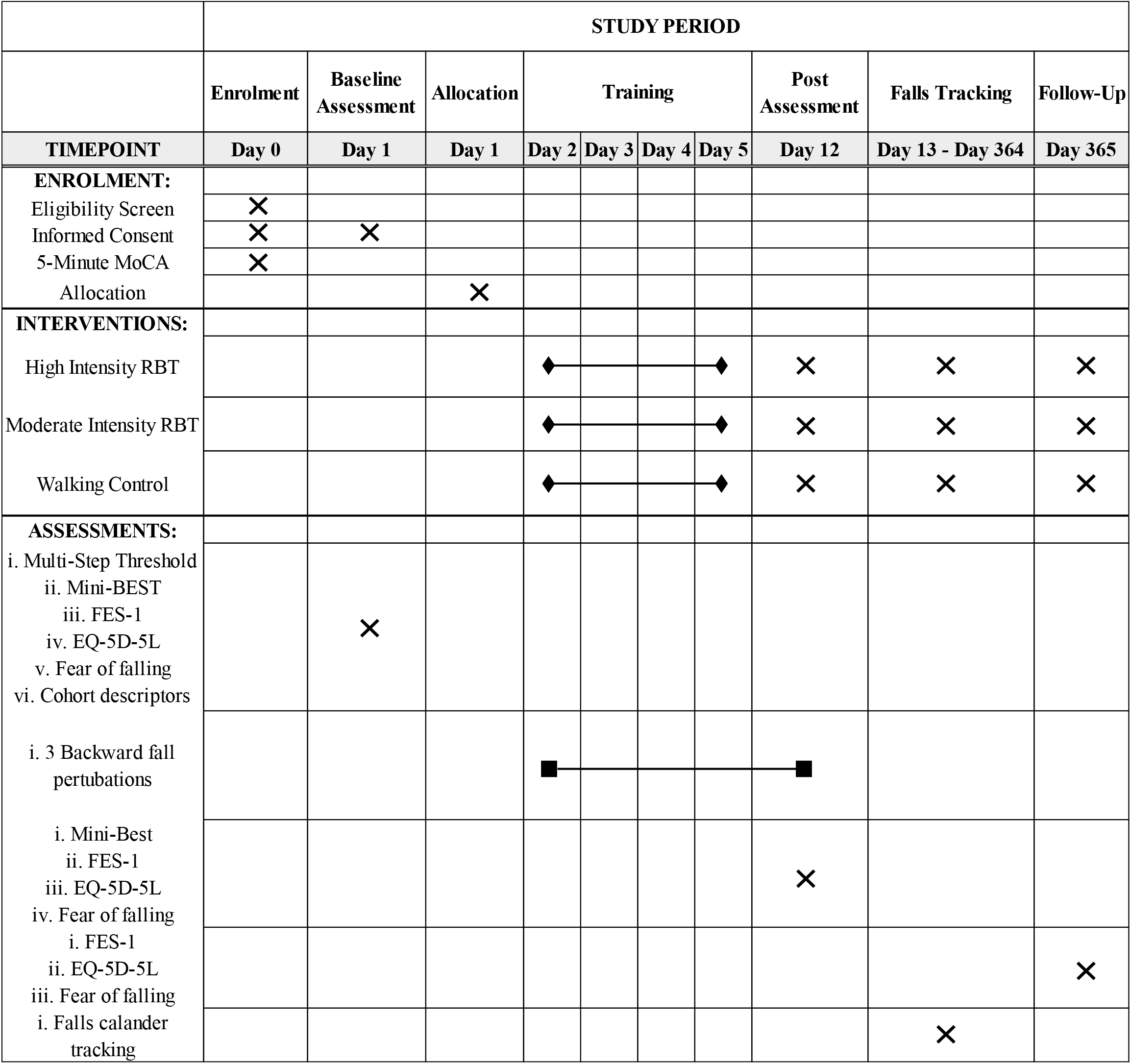
SPIRIT for the schedule of enrolment, interventions, and assessments.

## 2. METHODS: PARTICIPANTS, INTERVENTIONS, OUTCOMES

### 2.1 Study setting

The study will be conducted in a research laboratory at KITE, Toronto Rehabilitation Institute, University Health Network in Toronto, Ontario, Canada.

### 2.2 Eligibility criteria

We will recruit participants from in and around the greater Toronto area. Sixty-three adults (>20 years old) with chronic stroke (>6 months post-stroke) will be recruited. Participants will be excluded from participating in this study if they:

1. are unable to stand independently without upper-limb support for >30 seconds and/or walk independently (without a gait aid) for ≥10 metres;
2. have another neurological condition that could affect balance control (e.g., Parkinson’s disease);
3. have cognitive impairment (5-Minute Montreal Cognitive Assessment^24^ score <12)or severe language or communication difficulties affecting understanding instructions;^25^
4. have contraindications to RBT, such as osteoporosis, activity restrictions due to cardiac event/surgery, or severe spasticity in the lower extremity;^16^ and/or
5. are currently attending in-or out-patient physiotherapy or supervised exercise.

### 2.3 Interventions

The training will occur using a 6m x 3m motion platform, with maximum acceleration, velocity, and displacement of 10 m/s^2^, of 2 m/s, and 2 m, respectively. All balance perturbations will be a square waveform, consisting of a 300 ms acceleration followed by either a 300 ms (‘standard’ waveform) or 600 ms (‘extended’ waveform) deceleration.^26^ The two waveforms will be used to prevent participants from learning to use the deceleration phase to aid balance recovery.^27^ Participants will wear a safety harness attached to a robotic gantry overhead to allow free movement and prevent falls to the floor.

During the pre-training assessment, we will determine participants’ multi-step threshold (MST) in the forward, left, and right directions (Figure 2). The multi-step threshold is defined as the highest perturbation magnitude for which participants can recover balance with a single step. The multi-step threshold will be determined using a staircase procedure.^28^ Participants will initially experience one perturbation in each direction with peak acceleration of 0.50 m/s^2^. Participants will be instructed to do whatever they need to do to recover balance but, if they need to take a step, to minimize the number of steps taken. If participants do not step to recover their balance after the first perturbation the intensity will be increased by 1.00 m/s^2^ until they execute a single step and then the intensity will be increased in 0.50 m/s^2^ increments. If participants only take a single step to recover balance after the first perturbation, the intensity will be increased in 0.50 m/s^2^ increments. The intensity will continue to increase until the participants execute a multi-step response (3 or more steps if a side step sequence^29^ – that is, a medial step followed by a lateral step – is executed, 2 or more steps for other step patterns) to recover their balance.^29^ Once this occurs, the intensity will then be reduced in 0.25 m/s^2^ increments until the participant only takes one step (first reversal). The intensity will be increased, and then decreased, in 0.25 m/s^2^ increments, reversing when the participant’s step response changes (i.e., single-or multi-step reaction) until there are four reversals (Figure 2).

**Figure 2.**
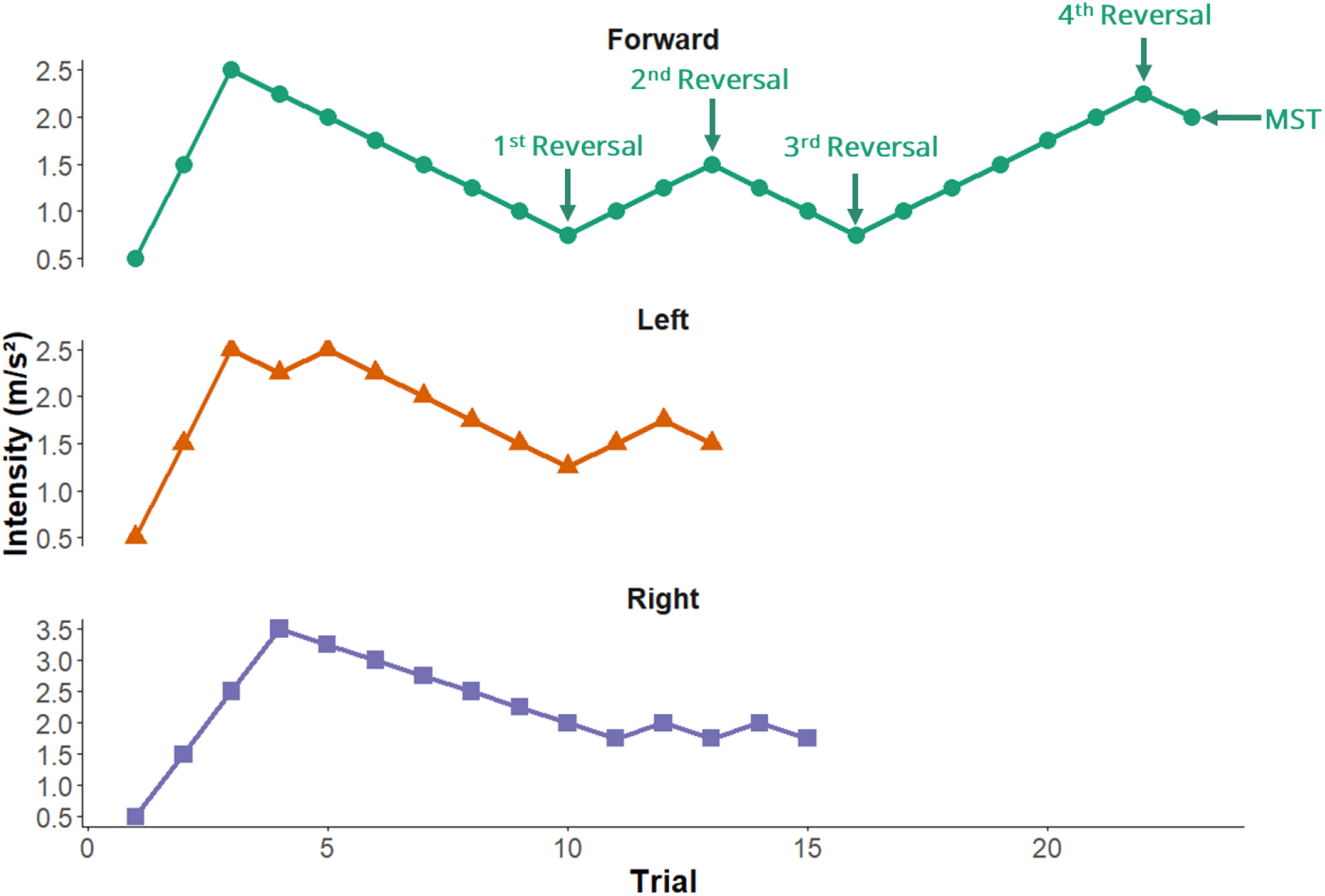
Example of the multi-step threshold for each direction, forward (blue), left (orange), right (green).

Training sessions will be conducted over four consecutive days. Each session will be scheduled for 1 hour and will be supervised by a licensed physiotherapist. Participants assigned to the high- and moderate-intensity RBT groups will experience 36 multi-directional (left-, right-, and forward ‘fall’) perturbations in each training session, presented in an unpredictable sequence and will be given 3 warm-up trials before each training session. The intensity of the warmup trials will depend on the training group each participant has been randomized into prior to the first session.

For participants in the moderate intensity group, 3 warm-up trials will begin at 66% of participants’ MST. If they maintain their balance without taking any steps in all three directions, the intensity for the remainder of the training session will be at their MST. However, if they take one step in all three directions, or a single step in only one lateral direction, they will remain at 66% of their MST for that training session. Should they take multiple steps in any one of the three directions, the intensity will be reduced by approximately 10%. In the high intensity group, the 3 warm-up trials will start at 100% of participants’ MST. If they take exactly one step in each of the three directions during these trials, the training intensity will continue at 150% of their MST for the remainder of the training session. If they take multiple steps in all three directions, or multiple steps in just one lateral direction, they will stay at 100% of their MST for the remainder of the training session. If participants’ in the high intensity group fall in any direction during the warm-up, the intensity will be reduced by 10% for the remainder of the training sessions.

Participants allocated to the high-intensity RBT group will complete perturbations at 150% of the direction-specific multi-step threshold; for example, for a multi-step threshold of 2 m/s^2^ the high intensity will be 3 m/s^2^. Participants assigned to the moderate-intensity RBT group will complete perturbations at the direction-specific multi-step threshold. If participants respond to the perturbations with a single step (high-intensity group) or no step (moderate-intensity group) in >50% of trials, the intensity will be increased by 25% in the following training session. Participants allocated to the walking control group will complete 36 walking trials on the platform in each training session. Rest breaks will be encouraged to minimize fatigue.

The number of perturbations completed in a session may be reduced, or sessions may be skipped/terminated, if participants’ report adverse outcomes (e.g., pain or fatigue). If participants are fearful of the training interventions the physiotherapist may decide to reduce the intensity regardless of the stepping responses to ensure the training is tolerable. All modifications to training will be made by the physiotherapist. To improve adherence to training, participants will be reimbursed for travel expenses. Participants will be asked not to change their regular exercise routine during the training portion of the study.

### 2.4 Outcomes

Outcome assessment will be completed pre-training, at the end of each training session, and one week after the last training session. We will also track falls in the participants for 12 months after the last retention visit.

#### 2.4.1 Primary outcomes

To assess the transfer of learning, participants will experience novel untrained (i.e., a ‘backward fall’) perturbations. To prevent participants from pre-planning their reactions, the backward-fall perturbations will be completed in a block of 9 trials with 6 low-magnitude forward, left and right perturbations at 25-50% of the direction-specific multi-step threshold, in an unpredictable order. The backward-fall perturbations will be completed at the forward-fall multi-step threshold. Because backward falls are more challenging than forward, backward-fall perturbations at the forward-fall multi-step threshold should evoke multi-step reactions.^28,30^

The primary outcome will be the number of steps taken to recover balance. Ineffective reactive steps will lead to additional steps to maintain balance,^31,32^ and multi-stepping is associated with an increased risk of falls in older adults.^12,33^ We will calculate the learning rate for each participant by fitting an exponential function to their data (average number of steps taken at each assessment time point), where *a* and *b* are constants, *t* is the assessment time point, and *x* is the adaptation rate.

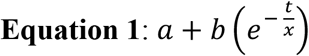

#### 2.2.2 Secondary outcomes: adverse outcome, mechanisms of improved reactive stepping, functional balance, and falls efficacy

Potential adverse events will be monitored, recorded, and reported by the physiotherapist. Examples of adverse events are injures, pain, fear, and anxiety related to the training interventions. The adverse events will be reported based on the ExHaRM protocol.^34^ Adverse events will be reported in our hospital’s health information system using the Common Terminology Criteria for Adverse Events (version 5.0; CTCAE).^35^ Any adverse events not included in the CTCAE will be documented separately in the training session log.

We will explore outcomes associated with reactive stepping ability: step initiation time (from perturbation onset to foot off), step execution time (from foot off to foot contact of the first step), step length (antero-posterior difference in foot position from foot off to foot contact), braking impulse (antero-posterior shear force generated over time on step contact), and mechanical margin of stability at foot contact.^36–38^ In addition, we will also administer the Mini-BEST, EQ-5D-5L, FES-I, fear of falling, and activity curtailment due to fear of falling will be assessed pre- and post-training.

#### 2.2.3 Participant characteristics

We will collect the following descriptors at baseline: age, sex, height, mass, time post-stroke, lesion location, pre-morbid medical history, prescription medications, the National Institutes of Health Stroke Scale (stroke severity),^39^ Chedoke-McMaster Stroke Assessment ^40^ foot and leg scores (motor impairment), and the Nielsen gender questionnaire.^41^

### 2.5 Participant timeline

We are currently recruiting for this study, and we estimate recruitment will end in October 2026, and data collection will end in October 2027. We expect to see the results of the study in November 2027.

Participants will be in the study for approximately 53 weeks. The first two weeks will involve outcome assessment and training, with a 1-year follow-up falls monitoring period completed at the end of the training sessions (Figure 1).

### 2.6 Sample size

The primary outcome is learning rate. The sample size for each group was estimated as λ/Δ,^42^ where λ was obtained from tables for 3 groups using probability of a Type 1 error (α) of 0.05, probability of a Type II error (β) of 0.2; therefore, λ of 9.64 was used. Additionally:

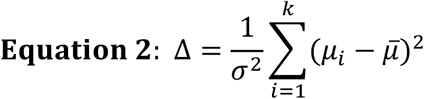

where *σ* is the standard deviation, *k* is the number of groups (3), *μ*_*i*_ is the mean for each group and 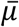 is the mean for all groups combined. From unpublished pilot data, we estimate a standard deviation of 9.7 for the learning rate. People with stroke typically require 3-4 steps to regain stability following the perturbation magnitudes used in the proposed study.^13,43^ Due to the mechanical challenges of the perturbation, at minimum 1 step is required to respond to the perturbation (i.e., young healthy adults would take 1 step to respond to the perturbation). Therefore, an improvement from 3-4 steps pre-training to 1-2 steps post-training is considered clinically meaningful, and a much larger effect size would likely not be possible. Previous studies have found that an improvement from taking on average, 3-4 steps pre-training to 1-2 steps post-training is feasible for people with chronic stroke.^13^ Simulating adaptation rates between these data points, we expect that adaptation rates of approximately 0.4 (*μ*_*1*_), 2 (*μ*_*2*_) and 10 (*μ*_*3*_) for the high-intensity, moderate-intensity, and walking control groups, respectively, would show meaningful differences between the three groups. Therefore, Δ was calculated as 0.56, giving a target sample size of 17 per group or 51 participants total. To account for a ∼20% rate of withdrawal,^12,44,45^ we will aim to recruit 21 participants per group, or 63 participants total. Enrolment will stop when 51 participants have completed the retention test, or 63 participants have been enrolled, whichever event occurs first.

### 2.7 Recruitment

Several strategies will be used to identify potential participants: 1) self-referral in response to advertisements; 2) referral from healthcare providers; or 3) patients who have indicated in their medical record that they are interested in being contacted about research. We will place advertisement posters in the community (i.e. poster boards, online, in community centres). For individuals responding to advertisements, the volunteer will be invited to contact the research team. For patients referred by healthcare providers at Toronto Rehab, initial contact may be made by a member of the research team (with the volunteer’s consent) or the volunteer may choose to contact the research team. Patients within our hospital can indicate in their medical record that they are interested in being contacted about research studies. We will identify these individuals within our hospital information system and will review their charts to pre-screen them using the eligibility criteria. Potentially eligible patients will be contacted by the research team using the patients’ preferred contact method, as indicated in their medical chart. To help to alleviate barriers to participating we will reimburse participants for travel expenses, and participants will receive a modest honorarium upon study completion.

## 3. METHODS: ASSIGNMENT OF INTERVENTIONS

### 3.1 Participant allocation

Participants will be assigned to one of the three intervention groups using blocked (block size 6 or 9) randomization, stratified by sex. The random allocation sequence will be computer-generated. The principal author (TNM) will screen potential participants for eligibility and enroll participants into the study. To ensure concealed allocation, the senior author (AM) or physiotherapist (CJD), who will not be involved in enrolling participants into the study, will assign participants to one of the three interventions groups.

### 3.2 Blinding (masking)

It is not possible to blind participants to group allocation, although participants in the moderate and high intensity groups may not realize which group they are in. The pre- and post-intervention tests will be administered by a blinded assessor. Responses to backward-fall perturbations will be administered at the end of each training session by non-blinded team members; however, the number of steps will be scored using video images by the blinded assessor. Post-processing to obtain secondary outcomes related to reactive stepping will also be completed by the blinded assessor. There is no foreseeable circumstance under which unblinding is permissible.

## 4. METHODS: DATA COLLECTION, MANAGEMENT, AND ANALYSIS

### 4.1 Data collection methods

All assessments and intervention sessions will take place in research laboratories at the Toronto Rehabilitation Institute. The mini-Balance Evaluation Systems Test, the Falls Efficacy Scale, and the Euroqol quality of life (EQ-5D-5L) measure^46–48^ will be obtained by a trained blinded research assistant. The mini-Balance Evaluation Systems test (mini-BEST), is a 14-item observational rating scale that assesses balance systems, including reactive balance control. The mini-BEST has good inter- (ICC=0.96) and intra-rater reliability (ICC=0.97) in chronic stroke.^49^ The 5-level EuroQoL health status measure (EQ-5D-5L) asks participants to rate their health in five dimensions (mobility, self-care, usual activities, pain/discomfort, and anxiety/depression) on a five-point scale and their overall health state on a scale from 0-100.The EQ-5D-5L has good-to-excellent test-retest reliability across several populations (ICC: 0.69-0.93).^50^ The FES-I asks participants to rate their concern about falling while performing 16 everyday tasks. The FES-I shows excellent test-retest reliability (ICC=0.97) in people with chronic stroke.^51^ Cohort descriptors will be obtained directly from participants, or from participants’ medical records. Our primary outcome (responses to untrained backward perturbations) will be obtained at the end of each training session and one week after the last training session to assess adaptation to the training using the custom moving platform. An accelerometer (Series 7523A, Dynamic Transducers and Systems, Chatsworth, California, USA) will be placed on the platform to record acceleration. We will collect 3D kinematic data using a markerless motion capture system (Theia Markerless, Inc., Kingston, ON), and use four force 1.5m x 1.5m plates placed on the platform (model BP11971197-2000, Advanced Mechanical Technology, Inc., Watertown, Massachusetts, USA) to record the participant’s forces and moments.

To assess fall incidence and injurious fall incidence, participants will be asked to report falls (“an event that results in a person coming to rest unintentionally on the ground or other lower level”^52^) for 12 months post-training. Participants will be provided with stamped, addressed postcards to mail to the research team every month for 1 year post-training. Postcards will contain a calendar, on which participants will record falls. The blinded research assistant will call participants who do not return the postcard to determine if any falls occurred. The research assistant will also contact participants reporting a fall to complete a short questionnaire determining the cause and consequences of the fall (e.g., injuries). This method is considered the ‘gold standard’ for fall reporting. ^53^

### 4.2 Data management

Baseline measurements recorded on physical copies (e.g., questionnaires, demographics) will undergo double data entry into an electronic database (e.g., Excel). Data collected during training sessions will be kept using a data collection sheet and data from the instrumentation collected (e.g., kinematic data, force plates files) will be stored in a computer file. All datasets will be maintained on secure, access-controlled servers.

### 4.3 Statistical methods

A one-way analysis of covariance (ANCOVA), using sex as a covariate, will compare the adaptation rate across the three intervention groups. ANCOVA, controlling for baseline values, will also compare post-intervention outcomes among groups, including average number of steps, step initiation time, step execution time, step length, braking impulse, margin of stability, mini-BEST scores, and FES-I scores. The alpha level will be set at 0.05, with adjustments for multiple comparisons using the Holm-Bonferroni method applied to the secondary outcome analyses. Adverse events will be categorized as present or absent and will be compared between groups using a chi-square test. This study is not powered for falls and quality of life outcomes; therefore, these data will be reported without inference so that these data can be used in meta-analyses and/or as pilot data for future studies. All participants will be analyzed as randomized; per protocol analysis may also be explored including only those participants who complete at least 80% of the intervention.

## 5. METHODS: MONITORING

### 5.1 Data monitoring

Given the low-risk nature of this intervention and the short duration of the reactive balance training interventions, a data monitoring committee is not necessary, as no substantial adverse events are anticipated. The intervention duration is short, limited to four sessions of 1 hour, and participant responses to the perturbations will be continuously and directly observed by a trained physiotherapist. Adverse events will be monitored by the principal investigator.

### 5.2 Harms

RBT has been conducted among individuals with stroke with few or no adverse events.^44,54–56^ Adverse events previously reported with RBT include delayed onset of muscle soreness, fatigue, and joint pain. There is also a risk of falling while participating in RBT or completing the reactive balance assessments. Participants will wear a safety harness attached to a secure point overhead in the event that they are unable to recover balance alone. The harness will ‘catch’ participants and prevent them from landing on the motion platform. To reduce the risk of injury, training will be overseen by an experienced licensed physiotherapist. In the event of injury while participating in this study, appropriate medical care will be provided.

### 5.3 Auditing

Study quality will be maintained through continuous monitoring by the principal investigator (AM) and co-investigator (EL). Additional auditing may be conducted by the Research Ethics Board, at their discretion.

## 6. ETHICS AND DISSEMINATION

### 6.1 Research ethics approval

This study was initially approved by the University Health Network Research Ethics Board (24-5322) on July 31^st^, 2024.

### 6.2 Protocol amendments

All protocol amendments will be submitted to the Research Ethics Board for approval. The research team will be informed of these amendments. If amendments impact enrolled research participants, they will be informed of the protocol changes and consent to continue the study will be obtained, if necessary.

### 6.3 Consent or assent

Participants will provide written informed consent prior to participation. The lead author (NM) will obtain consent from participants and will document the consent process.

### 6.4 Confidentiality

Several steps will be taken to ensure protection of personal health information. All information collected during this study, including the participant’s personal information, will be kept confidential and will not be shared with anyone outside the study unless required by law. Electronic data will be stored on secure UHN servers for 10 years. After 10 years the data will be deleted from the UHN servers. Electronic files containing patient names and contact information will be password protected and will be stored separately from study data. Hard copies of files containing de-identified data will be stored in locked cabinets and/or in offices that are locked when not occupied. Consent forms will be stored in locked cabinets/offices separately from other data. Only those individuals who require access to the data for the purpose of this study will be provided with the password to the file containing identifiers and/or the keys to the locked cabinet/office.

### 6.6 Access to data

No one outside of the research team will have access to the data, unless required as part of an internal audit by the Research Ethics Board.

### 6.7 Ancillary and post-trial care

If participants are hurt during the study, they will be referred for appropriate medical care. Participants will be people with chronic stroke, who have been discharged from any publicly funded post-stroke rehabilitation. As these participants do not receive rehabilitation services, post-trial care is not necessary.

### 6.8 Trial results

Study results will be shared with the academic community via publication in peer-reviewed journals and presentations at conferences. All authors who meet International Committee of Medical Journal Editors criteria for authorship will be included as authors in publications arriving from this research.^57^ No professional writers will be used. Study results will also be added to the trial registry. We will share results directly with physiotherapists through interactive workshops (e.g., at the Canadian Physiotherapy Association meeting). The results of the trial will be incorporated into our RBT toolkit.^58^ No plans are currently present for granting public access to the participant dataset.

## Data Availability

No datasets were generated or analyzed for the submitted manuscript.

## ACKNOWLEDGEMENTS

Funding for this project is provided by the Heart and Stroke Foundation of Canada, the Canada Brain Research Fund (CBRF), an innovative arrangement between the government of Canada (through Health Canada) and Brain Canada Foundation, and the Heart and Stroke Foundation of Canada (G-24-0038169).

## REFERENCES

1. Katan, M. & Luft, A. Global Burden of Stroke. Semin. Neurol. 38, 208–211 (2018).

2. Grefkes, C. & Fink, G. R. Recovery from stroke: current concepts and future perspectives. Neurol. Res. Pract. 2, 17 (2020).

3. Batchelor, F. A., Mackintosh, S. F., Said, C. M. & Hill, K. D. Falls after Stroke. Int. J. Stroke 7, 482–490 (2012).

4. Tyson, S. F., Hanley, M., Chillala, J., Selley, A. & Tallis, R. C. Balance Disability After Stroke. Phys. Ther. 86, 30–38 (2006).

5. Mansfield, A., Inness, E. L., Wong, J. S., Fraser, J. E. & McIlroy, W. E. Is Impaired Control of Reactive Stepping Related to Falls During Inpatient Stroke Rehabilitation? Neurorehabil. Neural Repair 27, 526–533 (2013).

6. Yu, H. et al. Effect of Cognitive Function on Balance and Posture Control after Stroke. Neural Plast. 2021, 1–6 (2021).

7. Andersson, Å.G., Kamwendo, K. & Appelros, P. Fear of falling in stroke patients: relationship with previous falls and functional characteristics. Int. J. Rehabil. Res. 31, 261–264 (2008).

8. Schmid, A. A. et al. Balance and Balance Self-Efficacy Are Associated With Activity and Participation After Stroke: A Cross-Sectional Study in People With Chronic Stroke. Arch. Phys. Med. Rehabil. 93, 1101–1107 (2012).

9. Lin, C.-L. Hsieh, S.-F. Hsiao, M.-H, J.-H. Predicting long-term care institution utilization among post-rehabilitation stroke patients in Taiwan: a medical centre-based study. Disabil. Rehabil. 23, 722–730 (2001).

10. Schmid, A. A. et al. Balance Is Associated with Quality of Life in Chronic Stroke. Top. Stroke Rehabil. 20, 340–346 (2013).

11. Carty, C. P. et al. Reactive stepping behaviour in response to forward loss of balance predicts future falls in community-dwelling older adults. Age Ageing 44, 109–115 (2015).

12. Mansfield, A., Peters, A. L., Liu, B. A. & Maki, B. E. Effect of a Perturbation-Based Balance Training Program on Compensatory Stepping and Grasping Reactions in Older Adults: A Randomized Controlled Trial. Phys. Ther. 90, 476–491 (2010).

13. Schinkel-Ivy, A., Huntley, A. H., Aqui, A. & Mansfield, A. Does Perturbation-Based Balance Training Improve Control of Reactive Stepping in Individuals with Chronic Stroke? J. Stroke Cerebrovasc. Dis. 28, 935–943 (2019).

14. Devasahayam, A. J. et al. The Effect of Reactive Balance Training on Falls in Daily Life: An Updated Systematic Review and Meta-Analysis. Phys. Ther. 103, pzac154 (2022).

15. Okubo, Y., Sturnieks, D. L., Brodie, M. A., Duran, L. & Lord, S. R. Effect of Reactive Balance Training Involving Repeated Slips and Trips on Balance Recovery Among Older Adults: A Blinded Randomized Controlled Trial. J. Gerontol. Ser. A 74, 1489–1496 (2019).

16. Mansfield, A., Danells, C. J., Inness, E. L., Musselman, K. & Salbach, N. M. A survey of Canadian healthcare professionals’ practices regarding reactive balance training. Physiother. Theory Pract. 37, 787–800 (2021).

17. Lippincott, Williams & Wilkins. ACSM, Guidelines for Exercise Testing and Prescription. (Philadelphia, 2010).

18. Wen, D. et al. Effects of different protocols of high intensity interval training for VO2max improvements in adults: A meta-analysis of randomised controlled trials. J. Sci. Med. Sport 22, 941–947 (2019).

19. Patel, P. & Bhatt, T. Adaptation to large-magnitude treadmill-based perturbations: improvements in reactive balance response. Physiol. Rep. 3, e12247 (2015).

20. Wang, Y., Wang, S., Lee, A.Pai, Y.-C. & Bhatt, T. Treadmill-gait slip training in community-dwelling older adults: mechanisms of immediate adaptation for a progressive ascending-mixed-intensity protocol. Exp. Brain Res. 237, 2305–2317 (2019).

21. Hróbjartsson, A. et al. SPIRIT 2025 explanation and elaboration: updated guideline for protocols of randomised trials. The BMJ 389, e081660 (2025).

22. Hoffmann, T. C. et al. Better reporting of interventions: template for intervention description and replication (TIDieR) checklist and guide. (2014) doi:10.1136/bmj.g1687.

23. Slade, S. C. et al. Consensus on Exercise Reporting Template (CERT): Modified Delphi Study.Phys. Ther. 96, 1514–1524 (2016).

24. Nasreddine, Z. S. MoCA Test: Validation of a five-minute telephone version. Alzheimers Dement.17, e057817 (2021).

25. Gagnon, G. et al. Correcting the MoCA for Education: Effect on Sensitivity. Can. J. Neurol. Sci. J. Can. Sci. Neurol. 40, 678–683 (2013).

26. Rajachandrakumar, R., Mann, J., Schinkel-Ivy, A. & Mansfield, A. Exploring the relationship between stability and variability of the centre of mass and centre of pressure. Gait Posture 63, 254– 259 (2018).

27. McIlroy, W. E. & Maki, B. E. The ‘deceleration response’ to transient perturbation of upright stance. Neurosci. Lett. 175, 13–16 (1994).

28. De Kam, D., Roelofs, J. M. B., Bruijnes, A. K. B. D., Geurts, A. C. H. & Weerdesteyn, V. The Next Step in Understanding Impaired Reactive Balance Control in People With Stroke: The Role of Defective Early Automatic Postural Responses. Neurorehabil. Neural Repair 31, 708–716 (2017).

29. Maki, B. E., Edmondstone, M. A. & McIlroy, W. E. Age-Related Differences in Laterally Directed Compensatory Stepping Behavior. J. Gerontol. A. Biol. Sci. Med. Sci. 55, M270–M277 (2000).

30. McIlroy, W. E. & Maki, B. E. Age-related Changes in Compensatory Stepping in Response to Unpredictable Perturbations. J. Gerontol. A. Biol. Sci. Med. Sci. 51A, M289–M296 (1996).

31. Wolfson, L. I., Whipple, R., Amerman, P. & Kleinberg, A. Stressing the Postural Response: A Quantitative Method for Testing Balance. J. Am. Geriatr. Soc. 34, 845–850 (1986).

32. Jacobs, J. V., Horak, F. B., Tran, K. & Nutt, J. G. An alternative clinical postural stability test for patients with Parkinson’s disease. J. Neurol. 253, 1404–1413 (2006).

33. Rogers, M. W. & Mille, M.-L. Lateral Stability and Falls in Older People: Exerc. Sport Sci. Rev. 31, 182–187 (2003).

34. Spence, R. R. et al. Practical suggestions for harms reporting in exercise oncology: the Exercise Harms Reporting Method (ExHaRM). BMJ Open 12, e067998 (2022).

35. Freites-Martinez, A., Santana, N., Arias-Santiago, S. & Viera, A. CTCAE versión 5.0. Evaluación de la gravedad de los eventos adversos dermatológicos de las terapias antineoplásicas. Actas Dermo-Sifiliográficas 112, 90–92 (2021).

36. Hsiao-Wecksler, E. T. Biomechanical and age-related differences in balance recovery using the tether-release method. J. Electromyogr. Kinesiol. 18, 179–187 (2008).

37. King, G. W., Luchies, C. W., Stylianou, A. P., Schiffman, J. M. & Thelen, D. G. Effects of step length on stepping responses used to arrest a forward fall. Gait Posture 22, 219–224 (2005).

38. Hof, A. L., Gazendam, M. G. J. & Sinke, W. E. The condition for dynamic stability. J. Biomech. 38, 1–8 (2005).

39. Goldstein, L. B., Bertels, C. & Davis, J. N. Interrater Reliability of the NIH Stroke Scale. Arch. Neurol. 46, 660–662 (1989).

40. Gowland, C. et al. Measuring physical impairment and disability with the Chedoke-McMaster Stroke Assessment. Stroke 24, 58–63 (1993).

41. Nielsen, M. W. et al. Gender-related variables for health research. Biol. Sex Differ. 12, 23 (2021).

42. Chow, S.-C., Shao, J., Wang, H. & Lokhnygina, Y. Sample Size Calculations in Clinical Research: Third Edition. (Chapman and Hall/CRC, Third edition. | Boca Raton: Taylor & Francis, 2017. | Series: Chapman & Hall/CRC biostatistics series | “A CRC title, part of the Taylor & Francis imprint, a member of the Taylor & Francis Group, the academic division of T&F Informa plc.”, 2017). doi:10.1201/9781315183084.

43. Schinkel-Ivy, A., Aqui, A., Danells, C. J. & Mansfield, A. Characterization of Reactions to Laterally Directed Perturbations in People With Chronic Stroke. Phys. Ther. 98, 585–594 (2018).

44. Mansfield, A. et al. Does perturbation-based balance training prevent falls among individuals with chronic stroke? A randomised controlled trial. BMJ Open 8, e021510 (2018).

45. Marigold, D. S. et al. Exercise Leads to Faster Postural Reflexes, Improved Balance and Mobility, and Fewer Falls in Older Persons with Chronic Stroke. J. Am. Geriatr. Soc. 53, 416–423 (2005).

46. Franchignoni, F., Horak, F., Godi, M., Nardone, A. & Giordano, A. Using psychometric techniques to improve the Balance Evaluation Systems Test: the mini-BESTest. J. Rehabil. Med. 42, 323–331 (2010).

47. Yardley, L. et al. Development and initial validation of the Falls Efficacy Scale-International (FES-I). Age Ageing 34, 614–619 (2005).

48. Herdman, M. et al. Development and preliminary testing of the new five-level version of EQ-5D (EQ-5D-5L). Qual. Life Res. 20, 1727–1736 (2011).

49. Tsang, C. S. L., Liao, L.-R., Chung, R. C. K. & Pang, M. Y. C. Psychometric properties of the Mini-Balance Evaluation Systems Test (Mini-BESTest) in community-dwelling individuals with chronic stroke. Phys. Ther. 93, 1102–1115 (2013).

50. Buchholz, I., Janssen, M. F., Kohlmann, T. & Feng, Y.-S. A Systematic Review of Studies Comparing the Measurement Properties of the Three-Level and Five-Level Versions of the EQ-5D. PharmacoEconomics 36, 645–661 (2018).

51. Hellström, K. & Lindmark, B. Fear of falling in patients with stroke: a reliability study. Clin. Rehabil. 13, 509–517 (1999).

52. Hyndman, D., Ashburn, A. & Stack, E. Fall events among people with stroke living in the community: circumstances of falls and characteristics of fallers. Arch. Phys. Med. Rehabil. 83, 165–170 (2002).

53. Myers, A. H., Baker, S. P., Van Natta, M. L., Abbey, H. & Robinson, E. G. Risk factors associated with falls and injuries among elderly institutionalized persons. Am. J. Epidemiol. 133, 1179–1190 (1991).

54. Mansfield, A. et al. Does Perturbation Training Prevent Falls after Discharge from Stroke Rehabilitation? A Prospective Cohort Study with Historical Control. J. Stroke Cerebrovasc. Dis. 26, 2174–2180 (2017).

55. Handelzalts, S. et al. Effects of Perturbation-Based Balance Training in Subacute Persons With Stroke: A Randomized Controlled Trial. Neurorehabil. Neural Repair 33, 213–224 (2019).

56. Bhatt, T., Dusane, S. & Patel, P. Does severity of motor impairment affect reactive adaptation and fall-risk in chronic stroke survivors? J. NeuroEngineering Rehabil. 16, 43 (2019).

57. ICMJE | Recommendations | Defining the Role of Authors and Contributors. https://www.icmje.org/recommendations/browse/roles-and-responsibilities/defining-the-role-of-authors-and-contributors.html.

58. Mansfield, A. et al. Implementing reactive balance training in rehabilitaton practice: a guide for healthcare professionals. (2021).

